# Evaluation of Eight Lateral Flow Tests for the detection of anti-SARS-CoV-2 antibodies in a vaccinated population

**DOI:** 10.1101/2022.04.04.22273232

**Authors:** Caitlin Greenland-Bews, Rachel. L. Byrne, Sophie. I. Owen, Rachel. L. Watkins, Daisy Bengey, Kate Buist, Karina Clerkin, Camille Escadafal, Lorna. S. Finch, Susan Gould, Emanuele Giorgi, Andy Hodgkinson, Larysa Mashenko, Darren Powell, Helen. R. Savage, Caitlin. R. Thompson, Lance Turtle, Jahanara Wardale, Dominic Wooding, Thomas Edwards, Ana Cubas Atienzar, Emily. R. Adams

**Affiliations:** Liverpool School of Tropical Medicine, Centre for Drugs and Diagnostics, Liverpool, UK; FIND, Foundation for Innovative New Diagnostics, Geneva, Switzerland; Department of Clinical Infection, Microbiology and Immunology, University of Liverpool, Liverpool; Mologic, Bedfordshire, UK; Lancaster University, Lancaster Medical School, Lancaster, UK; Biochemistry Department, Alder Hey Children’s Hospital, Liverpool, UK

**Keywords:** Antibody, COVID-19, Vaccination, Serology

## Abstract

With the distribution of COVID-19 vaccinations across the globe and the limited access in many countries, quick determination of an individual’s antibody status could be beneficial in allocating limited vaccine doses in low- and middle-income countries (LMIC). Antibody lateral flow tests (LFTs) have potential to address this need as a quick, point of care test, they also have a use case for identifying sero-negative individuals for novel therapeutics, and for epidemiology. Here we present a proof-of-concept evaluation of eight LFT brands using sera from 95 vaccinated individuals to determine sensitivity for detecting vaccination generated antibodies. All 95 (100%) participants tested positive for anti-spike antibodies by the chemiluminescent microparticle immunoassay (CMIA) reference standard post-dose two of their SARS-CoV-2 vaccine: BNT162b2 (Pfizer/BioNTech, n=60), AZD1222 (AstraZeneca, n=31), mRNA-1273 (Moderna, n=2) and Undeclared Vaccine Brand (n=2). Sensitivity increased from dose one to dose two in six out of eight LFTs with three tests achieving 100% sensitivity at dose two in detecting anti-spike antibodies. These tests are quick, low-cost point-of-care tools that can be used without prior training to establish antibody status and may prove valuable for allocating limited vaccine doses in LMICs to ensure those in at risk groups access the protection they need. Further investigation into their performance in vaccinated peoples is required before more widespread utilisation is considered.

## Introduction

In the ongoing COVID-19 pandemic, the rapid development and emergency use authorisation (EUA) of multiple COVID-19 vaccines (1–3) within the first year of the SARS-CoV-2 pandemic was an unprecedented achievement. Large-scale national vaccination programmes including booster shots are wide-spread in high income countries (4,5). This has sparked global discussion regarding vaccine equity (6,7) and the large disparity in the accessibility of COVID-19 vaccines between high- and low-income countries. Determination of antibody status would be beneficial in allocating vaccine doses according to serostatus. Model-based analysis has previously indicated that this strategy has potential to optimise the impact of a COVID vaccine dose (7,8). Furthermore, the use of monoclonal antibody therapies (mAbs) e.g., Ronapreve for treatment of COVID-19 patients requires that patients are seronegative to be eligible for therapy and therefore require rapid determination of antibody status before treatment can begin (9).

Determination of antibody titres to a specific pathogen is commonly achieved through enzyme linked immunosorbent assay (ELISA) or similar immunoassays, which are relatively accessible in high income countries, but less accessible in low- and middle-income countries (LMICs) (10). Lateral flow tests (LFTs) however are a quick, point of care test that require minimal prior training that could be scaled up for population wide screening for presence of anti-SARS-CoV-2 antibodies. These LFTs could be used to detect anti-SARS-CoV-2 antibodies quickly and determine suitability for COVID-19 vaccination or antibody therapy. Since the COVID-19 pandemic, an enormous number of manufacturers have developed LFTs which have entered the market without standardisation, although NIBSC now have available standards for anti-SARS-CoV-2 immunoglobulin. Still, there is minimal validation procedure for these tests, and to date the available data on these tests indicates variable performance (11–16). Lack of consistent methodology and reference standards make comparison of results between these studies difficult. Currently the World Health Organization (WHO) only recommends the use of these tests in research settings and states that more data are required on LFT performance to determine their suitability as a tool in the COVID-19 pandemic and global vaccination programme (17).

An evaluation of multiple brands of antibody LFTs is required in vaccinated individuals at multiple time points to accurately assess their performance compared to a sensitive reference standard. To this aim we have conducted an evaluation of eight commercially available LFTs with comparisons to an automated chemiluminescent microparticle immunoassay (CMIA) that is routinely used in clinical settings.

## Methods

### Study Design and Ethics

The NHS Research Ethics Committee (REC, UK) [REC reference:16/NW/0170] and the central Liverpool research ethics committee [Protocol Number: UoL001207] granted ethical approval for this work. The Integrated Research Application System (IRAS) Project ID is: 202413

Participants were recruited from the Liverpool School of Tropical Medicine and University of Liverpool staff networks as well as members of the public through social media outreach. Participants were recruited onto an existing study (The Human Immune Responses to Acute Viral Infections study (AVIS), 16/NW/0160). All participants gave written informed consent. Healthy individuals who had received or were due to receive their COVID-19 vaccination and were aged 18 years or over were recruited to the study. Individuals taking part in COVID vaccine trials were excluded from the study. Case record form (CRF) was completed by a trained member of staff to confirm eligibility. Participants were asked to provide a blood sample at days 21 (+/- 7 days), 42 (+/- 7 days) post dose one and two of their COVID-19 vaccine.

### Sample Collection and Processing

Venous blood (5ml, plain serum tube) samples were collected by trained health care workers and processed on the same day of collection. Briefly, venous blood samples were centrifuged at 1500g for 10 minutes and serum was aliquoted and stored at -20C° until testing.

### Lateral Flow Tests

LFTs were performed according to the instructions for use (IFU). Serum was allowed to thaw at room temperature for 15 minutes and vortexed for 5 seconds. According to individual IFU’s, 10-20µl of serum was added to the sample well and 2-3 drops of manufacturer specified buffer solution was added. Tests were run for 10-15 minutes, according to IFU, and read independently by two readers. Where there was a disagreement a third reader was used. Failed tests were repeated once. Characteristics of the of the tests used are shown in Table 1. When no details on the antigen composition were provided in the IFU, the company was approached for further information. Although all tests used in this study detect both IgM and IgG antibodies, IgG was the focus of this investigation and results from IgM are not included.

**Table 1:** Summary of Lateral Flow Test details including antigen, sample requirements and running time. RBD; receptor binding domain

|  |  |  |  |  |
| --- | --- | --- | --- | --- |
| LFT Brand | Antigen Used | Sample Volume (µl) | Buffer Volume (µl or drops) | Time to read (mins) |
| **WANTAI SARS-CoV-2 Ab Rapid Test (Beijing Wantai Biological Pharmacy) (Wantai)** | Spike-RBD | 10 | 2 drops | 15 |
| **Onsite COVID-19 IgG/IgM Rapid Test (CTK Biotech)) (CTK)** | Spike | 10 | 2 drops | 15 |
| **COVID-19 Total Ab Device (Fortress Diagnostics LTd) (Fortress)** | Spike- RBD | 10 | 2 drops | 10-15 |
| **NowCheck COVID-19 IgM/IgG Test (Bionote Co., LTD.) (Bionote)** | Nucleoprotein | 20 | 3 drops | 10-15 |
| **Edinburgh Genetics COVID-19 Colloidal Gold Immunoassay Testing Kit, IgG/IgM Combined (Edinburgh Genetics)** | Nucleoprotein | 20 | 60µl | 10-15 |
| **Diagnostic Kit for SARS-CoV-2 IgM/IgG Antibody (Colloidal Gold) (Shanghai Kehua Bio-Engineering Co., Ltd.) (KHB)** | Nucleoprotein | 10 | 3 drops | 15 |
| **SARS-CoV-2 IgM/IgG Ab Rapid Test (Qingdao HIGHTOP Biotech Co., Ltd.) (Qingdao)** | Nucleoprotein and Spike | 10 | 2 drops | 15-20 |
| **P4DETECT COVID-19 IgM/IgG (PRIME4DIA Co., Ltd) (Prime4Dia)** | Nucleoprotein and Spike | 10 | 3 drops | 10-15 |

### Immunoassays

Samples were analysed by quantitative and semi-qualitative chemiluminescent microparticle immunoassay (CMIA) on the fully automated Alinity i system (Abbot, United States) as a reference standard. SARS-CoV-2 IgG II CMIA (Abbott, Ireland) was used to quantify anti-S-RBD IgG antibodies in serum samples. To distinguish between antibodies produced from natural infection from SARS-CoV-2 to those produced from vaccination the samples were also analysed using the SARS-CoV-2 IgG I assay (Abbott, Ireland), a semi-qualitative CMIA to detect anti-nucleocapsid IgG antibodies, a method that was utilised by Narasimhan *et al*. and found to be effective (18). When using these assays, individuals positive for anti-nucleocapsid IgG antibodies are considered to have been naturally infected with SARS-CoV-2 and will also test positive for anti-S-RBD IgG antibodies. If an individual gives a negative test result for anti-nucleocapsid IgG antibodies but a positive result for anti-S-RBD IgG antibodies, then this individual is assumed to have not had a natural infection and has antibodies generated in response to vaccination. Following manufacturer recommendations, results higher or equal to 50AU/ml when using the SARS-CoV-2 IgG II Quant assay were considered positive for anti-S-RBD IgG antibodies. Similarly, results higher than or equal to 1.4 S/C (Sample control index) when using the SARS-CoV-2 IgG I Qualitative assay were considered positive for anti-nucleocapsid IgG antibodies as per manufacturer’s instructions.

### Model Formulation

A binomial mixed effect model was designed to analyse the impact of the key variables on the sensitivity of the different LFTs and to determine parameters for the calculation of the sensitivities and confidence intervals of each LFT at each dose. Details of modelling methods can be found in supplementary materials.

### Statistical Analysis

We used binomial mixed models to account for the clustering arising from the administration of multiple tests on the same individuals. These models were used to assess the effect of the LFT brand and other factors on the risk of a positive test. Due to the small sample size, binomial mixed models allowed us to borrow strength information across all individuals and estimate the sensitivity of each test more reliably than the conventional approach based on simple proportions. More details on the binomial mixed models and a comparison with the standard approach for estimation of LFT sensitivity can be found in the supplementary materials. The statistical analysis was conducted in RStudio (Version: 2021.9.1.372).

## Results

A total of 95 participants were recruited and provided at least one blood sample post dose one or two. A total of 89 participants provided a sample post dose one and 69 provided a sample post dose two with 63 participants providing a sample after both dose one and two. Of the 95 participants, 63 (66.3%) were female with a mean age of 39 years. CMIA analysis was conducted on all samples and showed that seven (10.1%) individuals tested positive for anti-nucleoprotein antibodies post-dose one and six (8.7%) tested positive post-dose two. The decrease in positivity is due to an individual not providing a sample post-dose 2 rather than loss of anti-nucleoprotein antibodies between doses. Of the seven participants that tested positive, five had previously reported a positive PCR test prior to enrolment with the study. CMIA also found 88 (98.8%) samples tested positive for anti-S-RBD antibodies post dose 1 and 69 (100%) post dose 2.

### Sensitivities

Point estimates of sensitivity from the binomial mixed effect model and the standard percentage calculation were largely comparable and results from both are summarised in table 2. For the remainder of the results, sensitivities quoted are from the model calculated values.

**Table 2:**
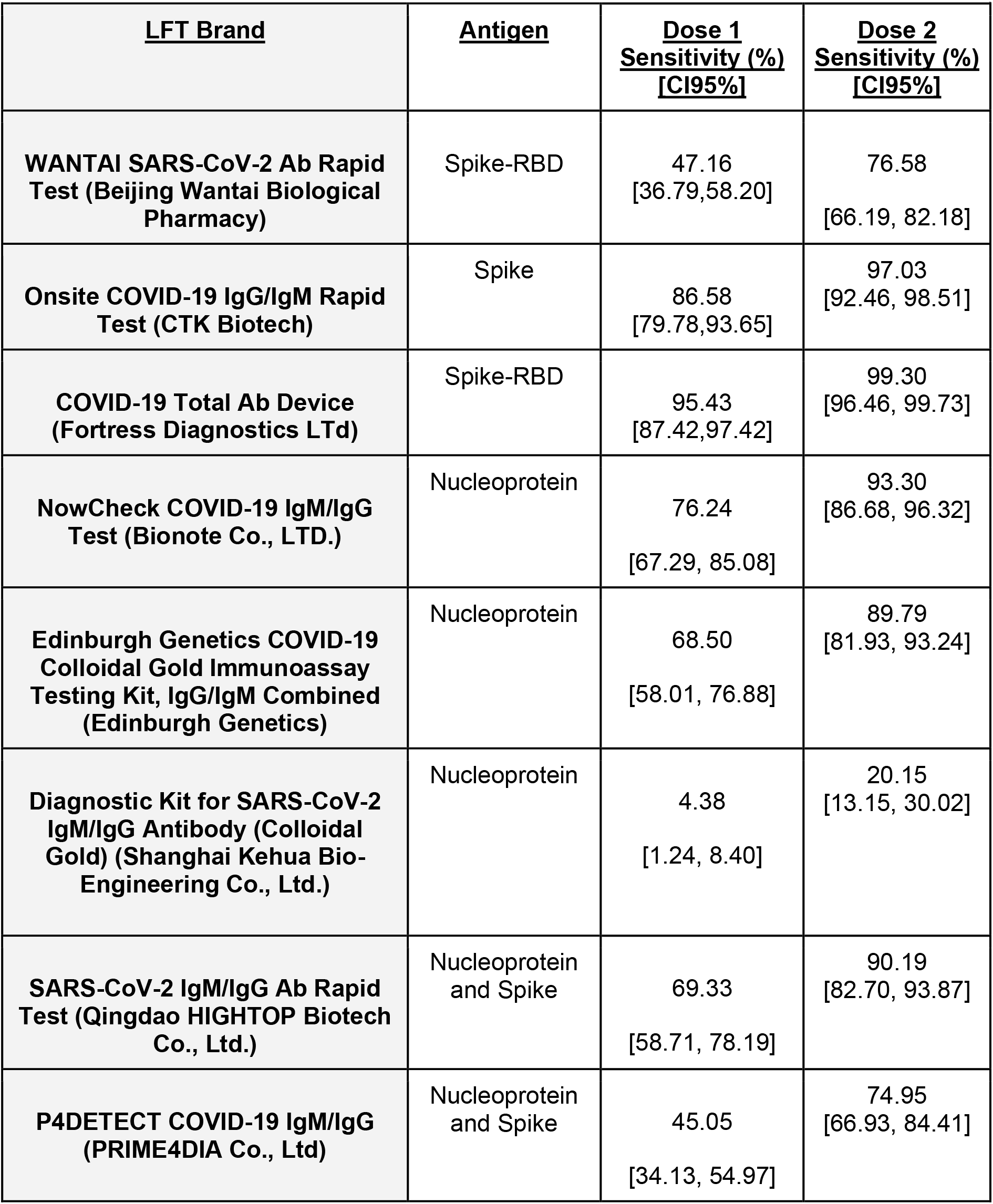
Point estimates and 95% confidence intervals from the lateral flow test (LFT) sensitivity obtained from the fitted binomial mixed model, for each brand at Dose 1 and 2.

Sensitivity for dose 1 ranged from 4.38% [CI95% 1.24,8.40] for KHB to 95.43% [87.42, 97.42] for Fortress. For dose 2, sensitivities ranged from 20.15% [13.15,30.02] for KHB to 99.30% [96.46,99.73] for Fortress.

Six out of eight LFTs showed a statistically significant increase in sensitivity estimates from dose 1 to dose 2. This may be indicative of higher antibody titres following a second vaccine dose making it easier to detect antibodies on LFT. Fortress did not have a statistically significant increase in sensitivity however had already achieved the highest sensitivity of all eight brands at dose 1.

Sensitivities of the tests when focussing on target antigen was varied. Three tests used nucleoprotein, two used both nucleoprotein and spike and three used spike alone, with two specifying the receptor binding domain (RBD) (Table 2). The tests that achieved the highest sensitivities post dose 2, CTK, Fortress and Bionote, all used different antigens of spike, spike-RBD and nucleoprotein respectively. The LFT with the lowest sensitivity was KHB which used nucleoprotein antigen.

### Impact of Variables on Test Result (Mixed Effect Model Analysis)

Our mixed effect model found that vaccine brand and day of sampling (Day 21 vs day 42) had no significant effect on overall test result and were therefore removed from the model analysis (supplementary materials, Table S1). Dose had a significant, positive effect on positivity rate with more positive results being detected after dose 2 compared to dose 1 (Table S2).

## Discussion

In this study we evaluated eight LFTs using sera from 95 vaccinated individuals, post-dose 1 and 2, to determine their sensitivity in detecting IgG antibodies specific to SARS-CoV-2 spike-RBD. We detected large variability in the sensitivities of these tests at different timepoints with Fortress having the highest sensitivity out of the eight tests evaluated, although specificity has not been considered in this study.

Overall, these results show LFTs can detect anti-S-RBD antibodies in vaccinated individuals and sensitivity increased with post-dose 2 samples. Sensitivity varies across the different brands and different antigens used. Fortress was the highest performing test in our vaccinated cohort and has also shown high sensitivity and specificity in other studies evaluating infected individuals(12,19,20) and is being implemented in a seroprevalence study in the UK(12,19). It was difficult to determine the impact the antigen in each test had on sensitivity and more information on the antigens from each brand would be beneficial for future evaluations.

Binomial mixed model analysis found that the test results were not significantly impacted by day of sample collection (Day 21 or 42 post vaccine dose) which is consistent with findings indicating IgG antibodies are detectable between 21 and 60 days after vaccination (21,22). Similarly, vaccine brand did not significantly impact test results, both these findings highlight that wider testing could be flexible without compromising sensitivity.

Future work should include evaluation of these tests in LMIC populations to build results for use in allocating vaccine priorities, and correlation studies to determine if a positive antibody LFT result is conducive to neutralising capacity in vaccinated individuals and if there is correlation between LFT line strength and protective antibody response. A recent review found imperfect correlation between presence of IgG and neutralising antibodies(23) however more investigation is required in this area.

LFTs have the potential to be a valuable, point of care tool to aid in the equitable distribution of vaccine doses according to existing antibody status and determine eligibility to life-saving monoclonal-antibody therapies. Our study has provided an evaluation of multiple brands of LFT in vaccinated people across multiple timepoints and the variation observed in our study and other evaluations(11–15) highlights the importance for robust evaluation methods and standardisation to be implemented.

## Data Availability

All data produced in the present study are available upon reasonable request to the corresponding authors

## Acknowledgements

This study received funding from the UK Research Council through a PhD scholarship from the MRC Doctoral Training Partnership to CGB. LT is supported by a Wellcome Trust fellowship [205228/Z/16/Z]. For the purpose of Open Access, the authors have applied a CC BY public copyright licence to any Author Accepted Manuscript version arising from this submission. LT, TF and EA are supported by the U.S. Food and Drug Administration Medical Countermeasures Initiative contract 75F40120C00085 and by the National Institute for Health Research Health Protection Research Unit (HPRU) in Emerging and Zoonotic Infections (NIHR200907) at University of Liverpool in partnership with Public Health England (PHE), in collaboration with Liverpool School of Tropical Medicine and the University of Oxford.

The views expressed are those of the author(s) and not necessarily those of the NHS, the NIHR, the Department of Health or Public Health England.

## Supplementary Materials

Let *Y*_*ij*_denote the test outcome for the i-th individual (1=“positive”, 0=“negative”) using the j-th brand. We then assume that the*Y*_*ij*_conditionally on a individual-level random effect, *Z*_*i*_, follows mutually independent Bernoulli variables with probability *p*_*ij*_for a positive test result (i.e. *Y*_*ij*_ = 1), such that

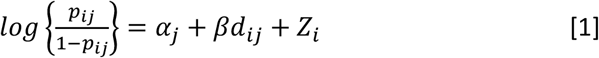

where *α*_*j*_is a brand-specific intercept and *d*_*ij*_is the number of the dose with associated regression coefficient β. Finally, we use *σ*^2^to denote the variance of *Z*_*i*_.

We carry out parameter estimation of the model in [1] using the Gaussian quadrature methods implemented in the glmer function of the lme4 (24) package in R.

We compute the sensitivity of a test as follows. Let *Y*_*s*_denote the outcome test for the reference test, and *Y*_*k*_the outcome of the test from any other brand. Let *P*[*A* | *B*] denote the probability of event A, given we have observed event B; the sensitivity of the k-th test, based on the model in [1], is then defined as

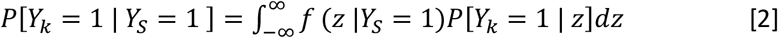

where *f*(*z* |*Y*_*s*_ = 1) is the density function of the individual-level random effects conditioned to having observed a positive reference test and *P*[*Y*_*k*_ = 1 | *z*]is the probability of a positive test based on the k-th brand, as defined in [1]. To compute the integral in [2] we use a quadrature approach to carry out numerical integration.

To compute the confidence intervals of the sensitivity estimate in [2], we proceed as follows.

1. We simulate 1000 samples for βand *σ*^2^ from a Multivariate Gassian distribution with mean given by the maximum likelihood estimates of βand *σ*^2^, and covariance matrix given by the inverse of the negative of the observed Fisher information.
2. For each of the 1000 samples simulated in the previous step, we compute the corresponding estimates of the sensitivity according to [2].
3. Using the simulated sensitivity values from the previous step, we compute the 0.025 and 0.975 quantiles to obtain a 95% confidence interval.

**Table S1.** Maximum likelihood estimates of the model parameters in equation [1].

| Parameter | Estimate | Confidence interval |
| --- | --- | --- |
| $\alpha_{ij}$ | | |
| CTK | 4.560 | (3.278, 5.842) |
| EdGen | 2.022 | (0.990, 3.053) |
| Fortress | 6.941 | (5.045, 8.837) |
| Hightop | 2.117 | (1.079, 3.154) |
| KHB | -6.559 | (-8.007, -5.111) |
| NowCheck | 2.962 | (1.863, 4.061) |
| NPRef | -8.064 | (-9.808, -6.319) |
| P4D | -0.419 | (-1.381, 0.544) |
| SpikeRef | 8.815 | (5.722, 11.908) |
| Wantai | -0.205 | (-1.168, 0.758) |
| $\beta$ | 3.169 | (2.483, 3.854) |
| $\sigma^2$ | 13.484 | (8.470, 21.686) |

## Validation of Binomial mixed model point estimates

Point estimates of LFT sensitivity from the binomial mixed model analysis were validated against proportional sensitivity that was calculated in Excel 2019 (Microsoft 365). Results from this validation are shown in Table S2 and S3

**Table S2:** Point estimates and 95% confidence intervals from the lateral flow test (LFT) sensitivity obtained from the fitted Binomial mixed model against proportional sensitivity, for each brand at Dose 1.

| **<u>LFT Brand</u>** | **<u>Antigen</u>** | **<u>Model Sensitivity (%)</u>**<br>**<u>[CI95%]</u>** | **<u>Proportional Sensitivity (%)</u>**<br>**<u>[CI95%]</u>** |
| --- | --- | --- | --- |
| **WANTAI SARS-CoV-2 Ab Rapid Test (Beijing Wantai Biological Pharmacy)** | Spike-RBD | 47.16<br>[36.79,58.20] | 40.91<br>[30.54,51.91] |
| **Onsite COVID-19 IgG/IgM Rapid Test (CTK Biotech)** | Spike | 86.58<br>[79.78,93.65] | 88.64<br>[80.09, 94.41] |
| **COVID-19 Total Ab Device (Fortress Diagnostics LTd)** | Spike-RBD | 95.43<br>[87.42,97.42] | 97.72<br>[92.3, 99.72] |
| **NowCheck COVID-19 IgM/IgG Test (Bionote Co., LTD.)** | Nucleoprotein | 76.24<br>[67.29, 85.08] | 76.14<br>[65.86, 84.58] |
| **Edinburgh Genetics COVID-19 Colloidal Gold Immunoassay Testing Kit, IgG/IgM Combined (Edinburgh Genetics)** | Nucleoprotein | 68.50 [58.01, 76.88] | 69.32 [58.58, 78.71] |
| **Diagnostic Kit for SARS-CoV-2 IgM/IgG Antibody (Colloidal Gold) (Shanghai Kehua Bio-Engineering Co., Ltd.)** | Nucleoprotein | 4.38<br>[1.24, 8.40] | 14.77<br>[8.11, 23.94] |
| **SARS-CoV-2 IgM/IgG Ab Rapid Test (Qingdao HIGHTOP Biotech Co., Ltd.)** | Nucleoprotein and Spike | 69.33<br>[58.71, 78.19] | 70.45<br>[59.78, 79.71] |
| **P4DETECT COVID-19 IgM/IgG (PRIME4DIA Co., Ltd)** | Nucleoprotein and Spike | 45.05<br>[34.13, 54.97] | 37.50<br>[27.40, 48.47] |

**Table S3:**
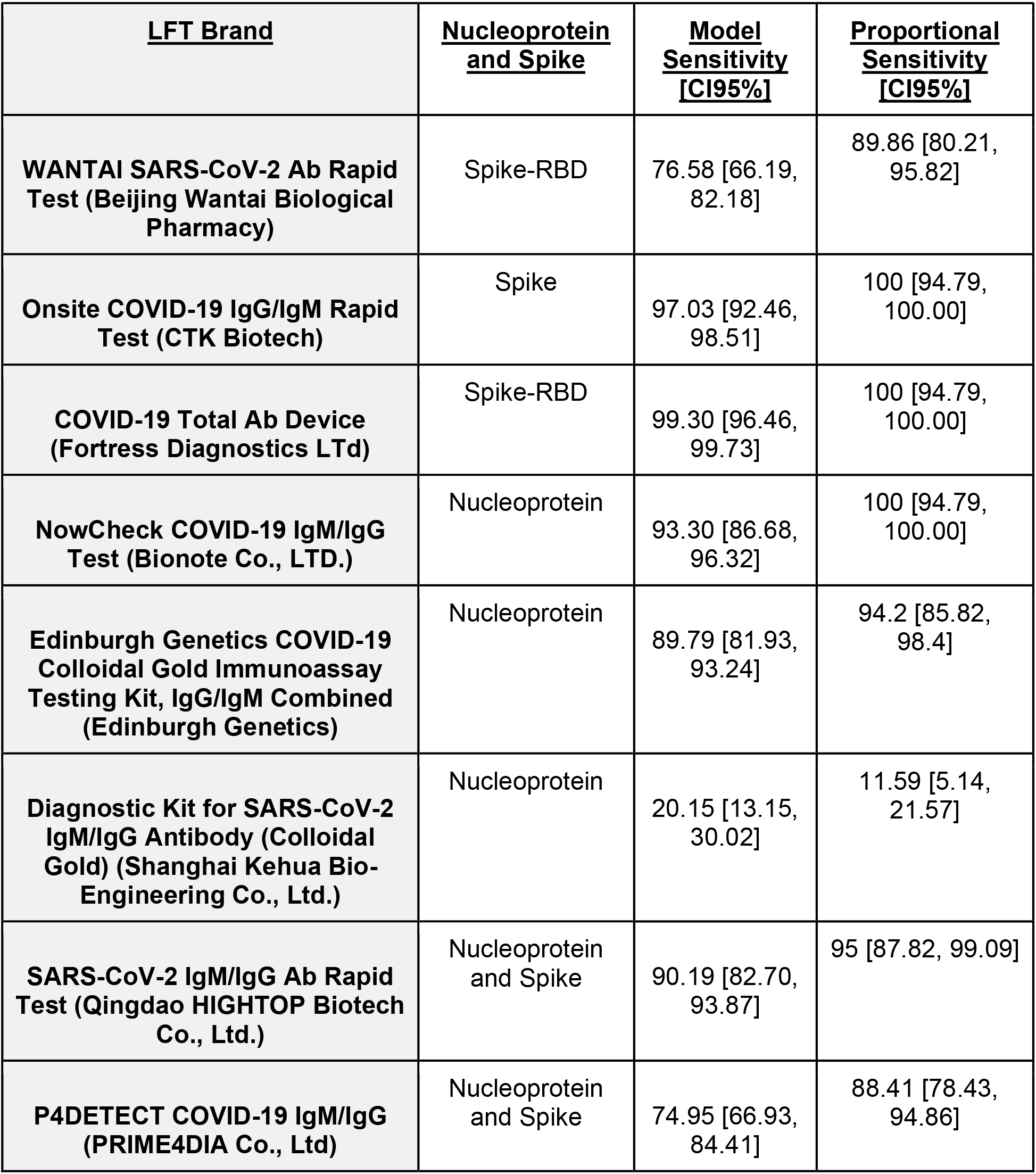
Validation of point estimates and 95% confidence intervals from the lateral flow test (LFT) sensitivity obtained from the fitted Binomial mixed model against proportional sensitivity, for each brand at Dose 2.

## References

1. Medicines and Healthcare products Regulatory Agency (MHRA). Regulatory approval of Spikevax (formerly COVID-19 Vaccine Moderna) - GOV.UK [Internet]. 2021 Jan [cited 2021 Dec 1]. Available from: https://www.gov.uk/government/publications/regulatory-approval-of-covid-19-vaccine-moderna

2. Medicines and Healthcare products Regulatory Agency (MHRA). Regulatory approval of Pfizer/BioNTech vaccine for COVID-19 - GOV.UK [Internet]. 2020 Dec [cited 2021 Dec 1]. Available from: https://www.gov.uk/government/publications/regulatory-approval-of-pfizer-biontech-vaccine-for-covid-19

3. Medicines and Healthcare products Regulatory Agency (MHRA). Regulatory approval of Vaxzevria (previously COVID-19 Vaccine AstraZeneca) - GOV.UK [Internet]. 2020 Dec [cited 2021 Dec 1]. Available from: https://www.gov.uk/government/publications/regulatory-approval-of-covid-19-vaccine-astrazeneca

4. UK Health Security Agency (UKHSA). COVID-19 vaccination: a guide to booster vaccination for individuals aged 40 years and over - GOV.UK [Internet]. 2021 Nov [cited 2021 Dec 1]. Available from: https://www.gov.uk/government/publications/covid-19-vaccination-booster-dose-resources/covid-19-vaccination-a-guide-to-booster-vaccination

5. Centers for Disease Control and Prevention (CDC). COVID-19 Vaccine Booster Shots | CDC [Internet]. 2021 [cited 2021 Dec 1]. Available from: https://www.cdc.gov/coronavirus/2019-ncov/vaccines/booster-shot.html

6. Adhanom Ghebreyesus T. Why There Should Be a Moratorium on COVID-19 Booster Shots Until Low-Income Countries Get Vaccinated. TIME. 2021 Aug 12;

7. Wise J. Covid-19: People who have had infection might only need one dose of mRNA vaccine. Available from: https://www.medrxiv.org/content/10.1101/2021.01.29.21250653v1

8. Bubar KM, Reinholt K, Kissler SM, Lipsitch M, Cobey S, Grad YH, et al. Model-informed COVID-19 vaccine prioritization strategies by age and serostatus. Science (1979) [Internet]. 2021 Feb 26 [cited 2021 Dec 1];371(6532):916–21. Available from: https://www.science.org

9. National Insitute for Health and Care Excellence (NICE). NICE updates managing COVID guideline with new monoclonal antibody recommendations | News and features | News | NICE. [cited 2022 Jan 25]; Available from: https://www.nice.org.uk/news/article/nice-updates-managing-covid-guideline-with-new-monoclonal-antibody-recommendations

10. Peeling RW, Wedderburn CJ, Garcia PJ, Boeras D, Fongwen N, Nkengasong J, et al. Serology testing in the COVID-19 pandemic response. The Lancet Infectious Diseases. 2020 Sep 1;20(9):e245–9.

11. Conklin SE, Martin K, Manabe YC, Schmidt HA, Miller J, Keruly M, et al. Evaluation of Serological SARS-CoV-2 Lateral Flow Assays for Rapid Point-of-Care Testing. J Clin Microbiol [Internet]. 2021 Feb 1 [cited 2021 Dec 1];59(2). Available from: https://pubmed.ncbi.nlm.nih.gov/33208477/

12. Flower B, Brown JC, Simmons B, Moshe M, Frise R, Penn R, et al. Clinical and laboratory evaluation of SARS-CoV-2 lateral flow assays for use in a national COVID-19 seroprevalence survey. Thorax. 2020 Dec 1;75(12):1082–8.

13. Heyming TW, Nugent D, Tongol A, Knudsen-Robbins C, Hoang J, Schomberg J, et al. Rapid antibody testing for SARS-CoV-2 vaccine response in pediatric healthcare workers. International Journal of Infectious Diseases. 2021 Dec;113:1–6.

14. Ragnesola B, Jin D, Lamb CC, Shaz BH, Hillyer CD, Luchsinger LL. COVID19 antibody detection using lateral flow assay tests in a cohort of convalescent plasma donors. BMC Research Notes. 2020 Aug 6;13(1).

15. van Elslande J, Houben E, Depypere M, Brackenier A, Desmet S, André E, et al. Diagnostic performance of seven rapid IgG/IgM antibody tests and the Euroimmun IgA/IgG ELISA in COVID-19 patients. Clinical Microbiology and Infection. 2020 Aug 1;26(8):1082–7.

16. National Institute for Biological Standards and Control (NIBSC). NIBSC - COVID-19-related research reagents [Internet]. [cited 2022 Mar 31]. Available from: https://www.nibsc.org/science_and_research/idd/cfar/covid-19_reagents.aspx

17. World Health Organisation. Laboratory testing strategy recommendations for COVID-19: Interim guidance. 2020 Mar.

18. Narasimhan M, Mahimainathan L, Araj E, Clark AE, Markantonis J, Green A, et al. Clinical evaluation of the abbott alinity sars-cov-2 spike-specific quantitative igg and igm assays among infected, recovered, and vaccinated groups. Journal of Clinical Microbiology [Internet]. 2021 Jun 1 [cited 2021 Dec 1];59(7). Available from: https://journals.asm.org/journal/jcm

19. Moshe M, Daunt A, Flower B, Simmons B, Brown JC, Frise R, et al. SARS-CoV-2 lateral flow assays for possible use in national covid-19 seroprevalence surveys (React 2): diagnostic accuracy study. [cited 2021 Dec 1]; Available from: http://dx.doi.org/10.1136/bmj.n423

20. Garrod G, Owen SI, Baillie JK, Baldwin L, Brown L, Byrne RL, et al. Comparative evaluation of ten lateral flow immunoassays to detect SARS-CoV-2 antibodies. Wellcome Open Research 2021 6:18 [Internet]. 2021 Feb 1 [cited 2021 Dec 1];6:18. Available from: https://wellcomeopenresearch.org/articles/6-18

21. Saluzzo F, Mantegani P, Poletti De Chaurand V, Quaresima V, Cugnata F, di Serio C, et al. SARS-CoV-2 Antibody Rapid Tests: Valuable Epidemiological Tools in Challenging Settings. Microbiology Spectrum. 2021 Oct 31;9(2).

22. Wei J, Stoesser N, Matthews PC, Ayoubkhani D, Studley R, Bell I, et al. Antibody responses to SARS-CoV-2 vaccines in 45,965 adults from the general population of the United Kingdom. Nature Microbiology 2021 6:9 [Internet]. 2021 Jul 21 [cited 2021 Dec 1];6(9):1140–9. Available from: https://www.nature.com/articles/s41564-021-00947-3

23. Savage HR, Santos VS, Edwards T, Giorgi E, Krishna S, Planche TD, et al. Prevalence of neutralising antibodies against SARS-CoV-2 in acute infection and convalescence: A systematic review and meta-analysis. PLoS Negl Trop Dis [Internet]. 2021 Jul 1 [cited 2021 Dec 1];15(7). Available from: https://pubmed.ncbi.nlm.nih.gov/34237072/

24. Bates D, Mächler M, Bolker BM, Walker SC. Fitting Linear Mixed-Effects Models Using lme4. Journal of Statistical Software [Internet]. 2015 Oct 7 [cited 2022 Jan 27];67(1):1–48. Available from: https://www.jstatsoft.org/index.php/jss/article/view/v067i01

